# Quantification and progress over time of specific antibodies against SARS-CoV-2 in breast milk of lactating women vaccinated with BNT162b2 Pfizer-BioNTech COVID-19 vaccine (LacCOVID)

**DOI:** 10.1101/2021.11.11.21266119

**Authors:** Erika Esteve-Palau, Araceli Gonzalez-Cuevas, M. Eugenia Guerrero, Clara Garcia-Terol, M. Carmen Alvarez, Geneva Garcia, Encarna Moreno, Francisco Medina, David Casadevall, Vicens Diaz-Brito

**Affiliations:** Department of Infectious Diseases, Parc Sanitari Sant Joan de Deu (Sant Boi, Barcelona, Spain); Department of Microbiology, Parc Sanitari Sant Joan de Deu (Sant Boi, Barcelona, Spain); Department of Obstetrics and Gynecology, Parc Sanitari Sant Joan de Deu (Sant Boi, Barcelona, Spain); Cancer Research Program, Hospital del Mar Research Institute, Barcelona, Spain

**Author notes:** Corresponding Author: Vicens Diaz-Brito, MD, PhD, MS, Department of Infectious Diseases, Parc Sanitari Sant Joan de Deu, 08830 Sant Boi, Barcelona, Spain.

## Abstract

**Importance:** To our knowledge, this is the first study to analyze long–term passage (6 months after immunization) of specific antibodies induced by BNT162b2 COVID-19 vaccine through breast milk.

**Objectives:** Main objective: to determine SARS-CoV-2 vaccine induced antibody levels in the breast milk of lactating women 4 weeks after mRNA BNT162b2 Pfizer-BioNTech COVID-19 complete vaccination.

Secondary objectives: to analyze SARS-CoV-2 antibody levels (breast milk and serum) at different time-points after vaccination, examine the correlation of SARS-CoV-2 antibody levels between serum and breast milk, describe adverse events related to vaccination (AErV) in both mothers and infants and determine the rate of COVID-19 infections.

**Design:** Prospective cohort study between February and September 2021.

**Setting:** Parc Sanitari Sant Joan de Déu, an urban hospital in Spain.

**Participants:** During our health worker vaccination campaign at our hospital between January and March 2, we recruited 33 lactating women vaccinated with BNT162b2 Pfizer-BioNTech COVID-19.

**Results:** A total of 33 volunteers were included in the study. The median (IQR) age of mothers was 38 (36-39) years and 15 (10-22) months for the infants.

Primary end-point: at 4 w after second dose median (IQR) IgG-S1 levels for serum–milk pairs were 12,478 (6,870-20,801) to 50.4 (24.3-104) arbitrary units (AU) per mL.

**Secondary end-points:** SARS-CoV-2 antibody levels at different time-points were (serum-milk): 519 (234-937) to 1 (0-2.9) AU/mL at 2w after first dose, 18,644 (9,923-29,264) to 78 (33.7-128) AU/mL at 2w, 4,094 (2,413-8,480) to 19.9 (10.8-51.9) AU/mL at 12w, and 1,350 (831-2,298) to 8.9 (7.8-31.5) at 24w after second dose.

We found a positive correlation of SARS-CoV-2 antibody levels between serum and breast milk (Pearson correlation coefficient 0.68).

No serious AErV were observed.

We found two (6%) COVID-19 vaccine breakthrough infections.

**Conclusions:** Pfizer-BioNTech COVID-19 vaccination is safe during breastfeeding and it transmits antibodies into breast milk with a positive correlation with serum levels, and both decrease over time in a 6-month follow-up. Infants of breastfeeding vaccinated women could be protected for at least six months after vaccination and serum determination of SARS-CoV-2 IgG-S1 could indicate the breastmilk levels of antibodies during this period.

## Introduction

Breastfeeding is one of the most efficacious means of preventing diseases and promoting health in both mothers and children^1^. Transfer of passive and active immunity through human milk is a key element in infant protection against infections^2,3^. In the neonatal period, newborns are exposed to a myriad of microorganisms, whose main entry point is through mucosal barriers, and infants initially have an immune system which is too immature to cope with pathogens.

Breast milk contributes to a significant reduction in infant morbidity and mortality when breastfeeding is performed exclusively in the first six months of life^4-9^. Apart from its nutritional richness, both colostrum and mature milk have a high content of immunoglobulins, proteins, lactoferrin and leukocytes, among other immunomodulatory factors, which makes it the first and best “vaccine” that the infant can receive in the first weeks and months of life^10,11^. In addition to nonspecific immunological compounds, specific antibodies against different infections are transmitted through breast milk, acquired from the mother’s previous contact with microorganisms or through the vaccines received against them^12,13^.

The COVID-19 pandemic has raised many questions among people who are breastfeeding, both because of the possibility of viral transmission to infants during breastfeeding and, with the approval of the SARS-CoV-2 vaccines, of the potential risks and benefits of vaccination in this specific population. Pregnant and breastfeeding women were excluded from all pre-marketing trials of anti-SARS-CoV-2 vaccines, so some doubts exist regarding its compatibility. In this regard, a meta-analysis of 48 studies with 183 infected women analyzed the rate of SARS-CoV-2 genome identification in breast milk, concluding that this was found in 5% of cases, associated mainly with mild cases of COVID-19 in breastfed infants^14^. However, other studies have observed that although SARS-CoV-2 RNA was found through PCR in the milk of infected women, these could not be detected in culture, suggesting that breast milk may not pose a risk of infection for the infant^15,16^. Different studies during the pandemic suggest that, far from posing a risk of infection to the infant, breast milk from infected mothers may be protective as it contains specific antibodies against SARS-CoV-2^14-17^.

More recently, several observational studies have also demonstrated the passage of postvaccine antibodies through breast milk in women vaccinated against COVID-19, mostly with mRNA-based vaccines^18-22^, but none showed long-term data. Thus, further research is needed to determine how long the antibodies are present in the breast milk of lactating mothers vaccinated against SARS-CoV-2.

Due to lack of knowledge in this field, our research group published the preliminary results in advance (1 month after mRNA vaccination), which demonstrated the passage of antibodies into breast milk^18^. Here we show the original research study with complete follow-up.

## Methods

We conducted a prospective cohort study between February and September 2021 at Parc Sanitari Sant Joan de Déu, an urban hospital in Spain, carried out according to the Strengthening the Reporting of Observational Studies in Epidemiology (STROBE) reporting guideline.

The primary end-point was to determine SARS-CoV-2 vaccine induced antibody levels in the breast milk of lactating women 4 weeks after mRNA BNT162b2 Pfizer-BioNTech COVID-19 complete vaccination. Secondary end-points were to examine the correlation of SARS-CoV-2 antibody levels between serum and breast milk, analyze SARS-CoV-2 antibody levels (breast milk and serum) at different time-points after vaccination, describe adverse events related to vaccination (AErV) in both mothers and infants and determine the rate of COVID-19 infections. The study was explained to all volunteers in detail, and all gave written informed consent prior to participating. The Sant Joan de Déu Research Foundation ethics committee approved this study.

Inclusion criteria were: lactating women older than 18 years who were vaccinated against SARS-CoV-2 with the BNT162b2 Pfizer-BioNTech COVID-19 vaccine during the vaccination campaign between January and March 2021. All were health workers.

Serum and breast milk samples were simultaneously taken from each participant at different time points after vaccination: time point 1 (two weeks after first dose), time point 2 (two weeks after second dose), time point 3 (four weeks after second dose), time point 4 (twelve weeks after second dose) and time point 5 (twenty-four weeks after second dose). Moreover, all participants underwent nasopharyngeal Panbio® Abbot COVID-19 antigen rapid test (Ag-RDT) at each time point.

When blood samples were obtained, they were centrifuged for 15 minutes at 3,500 rpm and processed to determine levels of immunoglobulin (Ig) G antibodies against the spike protein S1 subunit (IgG-S1) and against the nucleocapsid (IgG-NC) of SARS-CoV-2 (Architect, Abbott®). As vaccination does not induce nucleocapsid antibody response, any IgG-NC positive result was considered a prior infection.

The milk samples were centrifuged for 15 minutes at 3,500 rpm and after removing the fat layer with a pipette, the liquid layer of the milk was collected. Subsequently, we repeated the same process once more to determine IgG-S1 (Architect, Abbott®). Only in the case of obtaining positive IgG-NC in serum was determination carried out in milk.

No tests were performed on the infants. In cases of suspicion of COVID-19 infection or close contact, the infants were referred to pediatrics to perform a PCR SARS-CoV-2 assay to exclude COVID-19.

Variables related to parity and breastfeeding were collected, as well as the medical history of both mothers and infants. The participants were also questioned at each visit about COVID19 compatible symptoms in them or their infants and any adverse events related to the vaccine. Numeric variables were summarized using median and IQR (Q1-Q3). Correlation of serum and milk antibodies was analyzed using Pearson’s r. Statistical analyses were performed with R version 4.0.3 (R Project for Statistical Computing), and figures created using ggplot2 R package.

## Results

### Characteristics of participants and samples

A total of 33 volunteers were included in the study. The median (IQR) age of mothers was 38 (36-39) years and 15 (10-22) months for the infants at the time of the vaccination. All mother and infant characteristics are described in Table 1.

**Table 1.** Participants’ characteristics.

|  |  |
| --- | --- |
| Age | 38 (36-39) years |
| Delivery type | Natural vaginal delivery = 21 (64%); induced vaginal delivery = 9 (27%); elective caesarian section = 1 (3%); emergency caesarian section 2 = (6%). |
| Gestational age (last delivery) | 40 (38-40) weeks |
| Mother’s BMI* | 21 (19.4-24.3) kg/m<sup>2</sup> |
| Medical history | MTHFR** gene mutation = 1 (3%); Early puberty = 1 (3%); Intraepithelial neoplasia of cervix grade 3 = 1 (3%); Urethral stricture = 1 (3%); Leiden factor V = 1 (3%); Raynaud syndrome = 1 (3%); Celiac disease = 1 (3%); Autoimmune thyroiditis = 1 (3%); Hypothyroidism = 1 (3%); Tuberculosis in childhood = 1 (3%). |
| Infant’s gender | Female = 20 (60.6%); Male = 13 (39.4%) |
| Infant’s age (at the time of the vaccine) | 15 (10-22) months |
| Birth weight | 3,200 (3,010-3,570) grams |
| Breastfeeding | Exclusive breastfeeding first 6 months = 32 (97%); Mixed breastfeeding = 1 (3%) |
| Profession | Doctor = 9 (27.3%) [4 physicians, 3 surgeons, 2 psychiatrists]<br>Nurse = 7 (21.2%), Midwife = 4 (12.1%), Nursing assistant = 4 (12.1%)<br>Social worker = 3 (9.1%), Nutritionist = 2 (6.1%), Administrative = 2 (6.1%)<br>Pharmacist = 1 (3%), Laboratory technician = 1 (3%) |

\*BMI: body mass index; \*\*MTHFR: 5-methyltetrahydrofolate. n: number of patients; %: percentage of total number of patients (n=33). Values are n (%) or median (IQR).

We were able to recruit and obtain samples to determine the primary end-point, antibody titers at 4 weeks after complete vaccination, in all participants (*n*=33). However, we only achieved samples after the first dose of vaccine in 28 participants, and in 32 at two weeks after the second dose. In the three-month follow-up, two participants were lost to follow-up (one due to not being able to express milk, the other due to having weaned) and at six-month follow-up a total of 8 participants were lost (4 were unable to express milk, 2 weaned, and 2 failed to visit). Number of volunteers and samples at each time point are shown in Figure 1.

**Figure 1.**
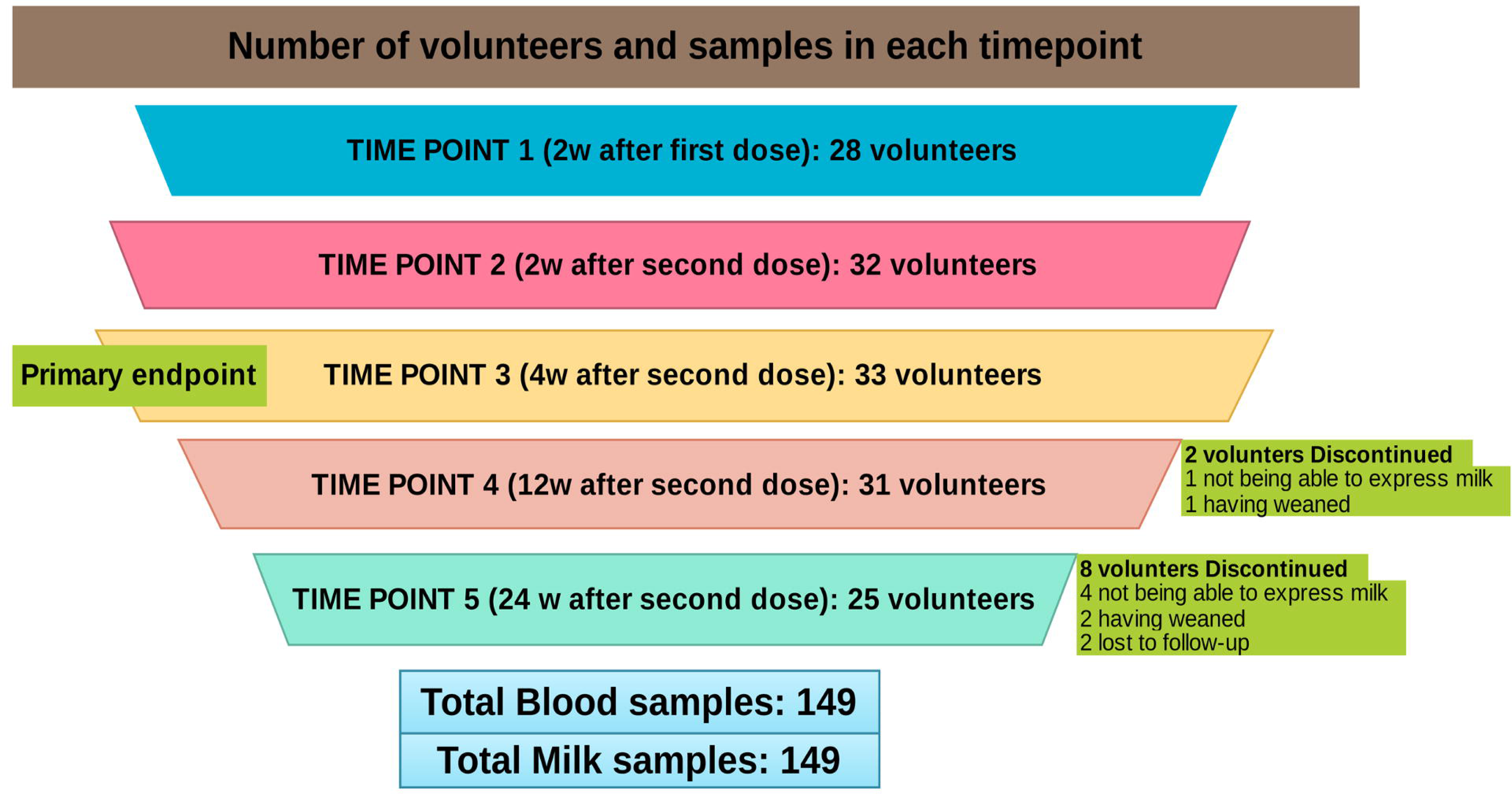
Number of volunteers and samples at each time point.

No participants had confirmed SARS-CoV-2 prior infection at the beginning of the study (blood tests for IgG-NC and nasopharyngeal Ag-RDT were all negative). We collected and analyzed 149 serum and 149 milk samples from the volunteers.

Samples from time point 1 were taken at a median (IQR) of 14 (13-16) days after the first dose, while time points 2 and 3 samples were taken at 14 (14-15) days and 28 (28-30) days after the second vaccine dose, respectively. Time point 4 samples were taken at 12-week follow-up 85 (84-88) days and time point 5 at 24-week follow-up 164 (161-174) days.

### SARS-CoV-2 vaccine induced antibodies and serum-breast milk correlation

At time point 3 (4 weeks after second dose, primary end-point), median (IQR) IgG-S1 levels for serum–milk pairs were 12,478 (6,870-20,801) to 50.4 (24.3-104) arbitrary units (AU) per mL. At different time-points after vaccination, IgG-S1 levels for serum–milk pairs were 519 (234-937) to 1 (0-2.9) AU/mL for time point 1, 18,644 (9,923-29,264) to 78 (33.7-128) AU/mL for time point 2, 4,094 (2,413-8,480) to 19.9 (10.8-51.9) AU/mL for time point 4, and 1,350 (831-2,298) to 8.9 (7.8-31.5) for time point 5. Although maximum levels of IgG-S1, in both serum and milk, were observed two weeks after the second dose of vaccine with a subsequently progressive parallel decrease, we found an increase in antibody levels in six-month follow-up samples in two participants related to a previous infection by SARS-CoV-2 (Figure 2). In these two cases, a positivization of IgG-NC was also observed in serum but not in breast milk.

**Figure 2.**
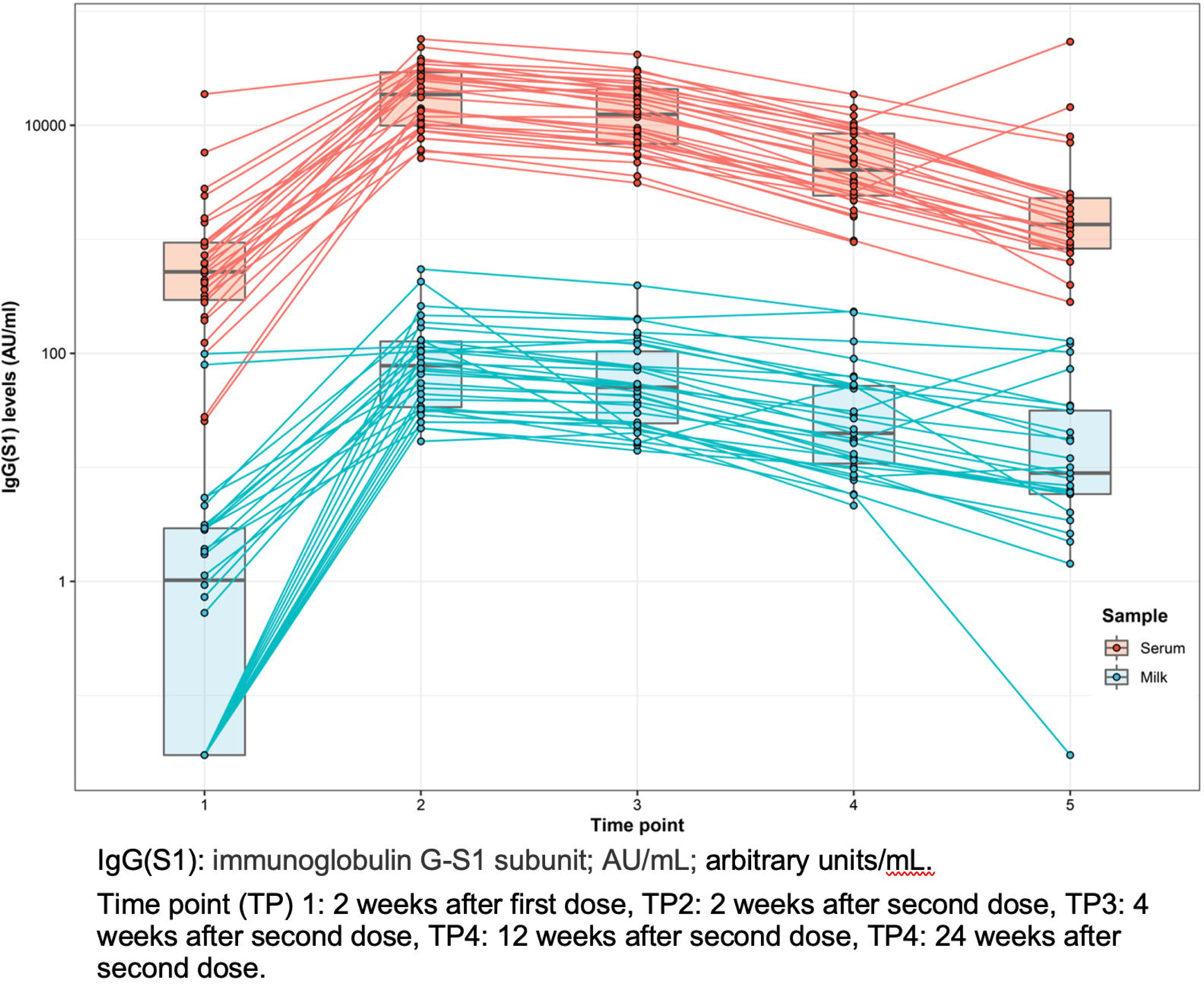
Progress over time of immunoglobulin G-S1 subunit levels in breast milk and serum of vaccinated participants across time.

We found that after the second dose, breast milk IgG-S1 levels increased and were positively associated with corresponding serum levels. Pearson correlation coefficient between breast milk and serum IgG-S1 levels was 0.68 (Figure 3).

**Figure 3.**
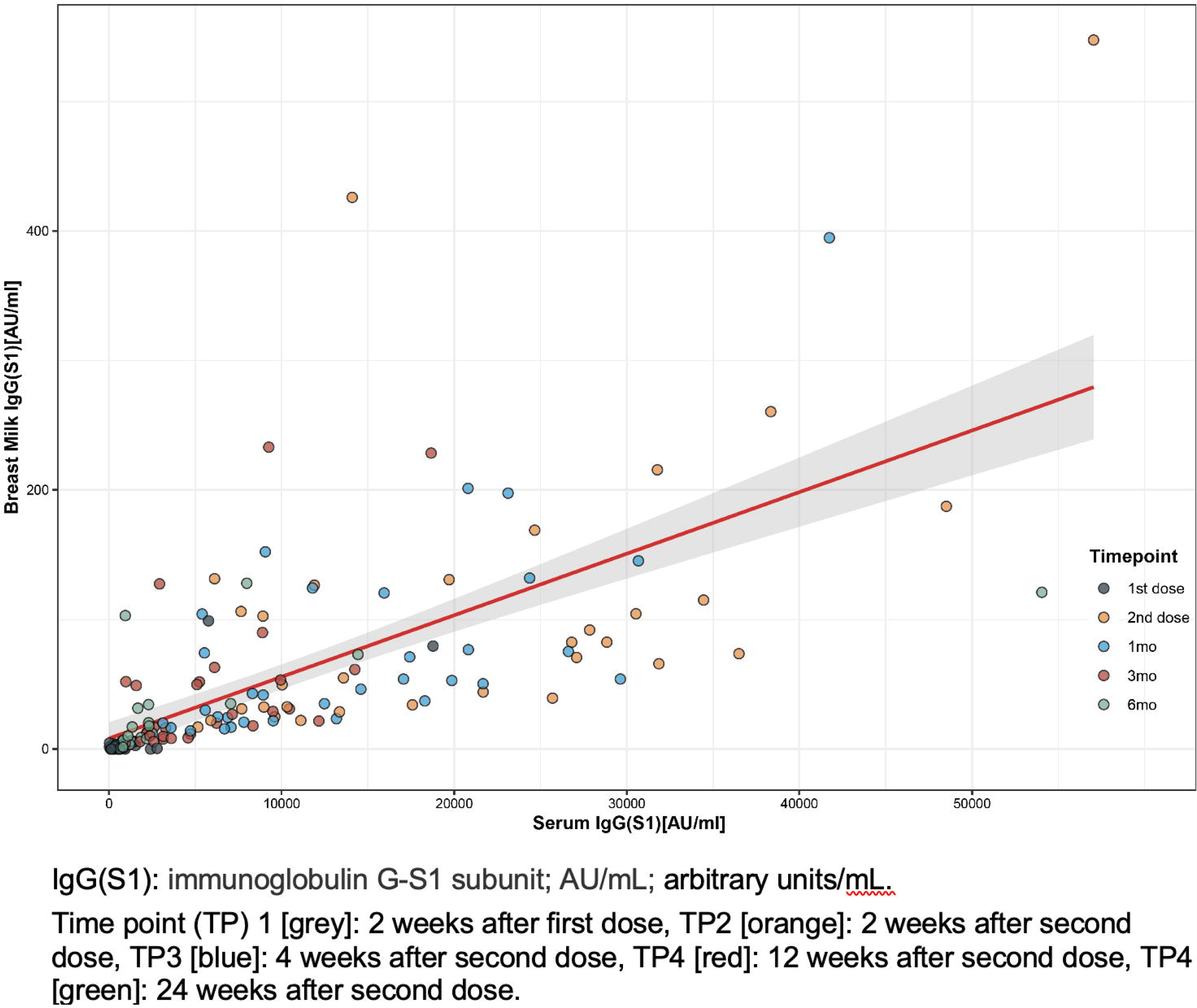
Correlation between immunoglobulin G-S1 subunit levels in serum and breast milk of vaccinated participants.

### Adverse events related to mRNA COVID-19 vaccination

The main AErV (mean of 2 doses) in our cohort were: pain at the injection site (95%), feverish feeling and/or confirmed fever (15%), general malaise (15%), headache (11%), arthromyalgia (9%), asthenia (6%), axillary adenopathy (5%), and other (<3%). All AErV are summarized in Table 2. No adverse events were observed in the infants. Number of accumulated AErV or having systemic symptoms related to the vaccination did not correlate with an increased humoral immunogenicity.

**Table 2.** Vaccine-related adverse events (AErV).

|  | *First dose\** | *Second dose\*\** |
| --- | --- | --- |
| *Local pain (mild-moderate)* | 32 (97%) | 31 (94%) |
| *Fever* | 3 (9%) | 7 (21%) |
| *General malaise* | 3 (9%) | 7 (21%) |
| *Headache* | 2 (6%) | 5 (15%) |
| *Arthromyalgia* | 1 (3%) | 5 (15%) |
| *Asthenia* | 1 (3%) | 3 (9%) |
| *Axillary adenopathy* | 2 (6%) | 1 (3%) |
| *Palmar erythema* | 1 (3%) | 1 (3%) |
| *Aphonia* | 1 (3%) | 0 |
| *Odynophagia* | 0 | 1 (3%) |
| *Nausea* | 0 | 1 (3%) |
| *Hypoglossal cyst* | 0 | 1 (3%) |

n: number of patients; %: percentage of total number of patients (n=33). Values are n (%).
\* First dose: AErV occurred during the 2 weeks after first dose vaccination.
\*\* Second dose: AErV occurred during the 2 weeks after complete vaccination.

### COVID-19 infections

During the study period, 2 participants had a COVID-19 vaccine breakthrough infection, one was completely asymptomatic, and the other had mild symptoms. In the first case, information on the contagion emerged from the analytical results of the six-month follow-up, in which a rebound in the level of IgG-S1 was observed in serum and milk compared to the previous sample at time point 4 (from 10,448 AU/ml to 54,045 AU/ml, and 31 AU/ml to 121 AU/ml, respectively), as well as a positivization of the IgG-NC antibodies in serum with an index of 2.45, and a negative value in milk. In this case, both the mother and her infant were asymptomatic between 3 and 6 months of follow-up. No diagnostic test was performed at the time of infection as it was asymptomatic and no diagnosis was required.

The second case was diagnosed with COVID-19 a week before the 6-month follow-up, with symptoms of mild upper respiratory infection and fever. Once the quarantine period was completed, new samples were taken (12 days after diagnosis and after a negative control PCR), showing a considerable increase in IgG-S1 in serum and milk compared to the previous determination at time point 4 (from 2,611 AU/ml to 14,425 AU/ml, and 17 AU/ml to 73 AU/ml, respectively) and an IgG-NC in serum with an index of 1.02 and negative in milk. The baby had fever and mild upper respiratory symptoms a few days later but no diagnostic test was performed since the mother had an obvious diagnosis and refused to perform a test on her child.

## Discussion

The current COVID-19 pandemic has raised multiple concerns for breastfeeding mothers. At first, due to the potential risk of infecting babies, breastfeeding was even contraindicated in infected women. After the approval of the SARS-CoV-2 vaccines, fears regarding breastfeeding centered on the potential harmful effects that these could have on babies. For this reason, and in the absence of scientific evidence, vaccination in lactating women was initially contraindicated in many settings, often leaving the choice of vaccination and/or maintaining breastfeeding in the hands of mothers.

Pregnant and breastfeeding women are often excluded from clinical trials with new drugs due to the fears and ethical dilemmas that these entail. This means that the safety of most drugs (including vaccines) is principally evaluated in post-marketing monitoring.

Given the lack of evidence regarding vaccination against COVID-19 in early 2021, Parc Sanitari Sant Joan de Déu promoted the LacCOVID study, which included lactating women who carry out their work on the front line and who decided to get vaccinated at the beginning of the vaccination campaign while continuing to breastfeed.

During the pandemic, different studies have shown that the infective virus is rarely transmitted through the milk of infected mothers^14-16^ and, instead, specific antibodies against the virus are transmitted to the babies through breast milk^18-22^. The passage of IgG specific antibodies into breast milk is also confirmed in our study in vaccinated women and, in addition, we were able to verify how these levels have a correlation with those observed in serum. These results are consistent with those observed in other recent research^21,22^. To date, there are no studies conducting long-term follow-up on the transmission of these antibodies to breast milk. To our knowledge, this is the first work with a six-month follow-up. In the three and six-month follow-up, we observed a progressive decrease in antibody levels in serum samples, which correlates with data from previous studies carried out in health care professionals vaccinated with the BNT162b2 vaccine^23,24^. Additionally, in our study, we observed that this decrease in serum antibody levels is associated with a parallel decrease in breast milk levels at 3 and 6 months, with the same serum-breast milk positive correlation. Therefore, it is reasonable to hypothesize that serum determination of SARS-CoV-2 IgG-S1 could indicate approximately the breastmilk levels of antibodies during the 6 months after vaccination.

Duration of transplacental immune protection against SARS-CoV-2 in infants of mRNA vaccinated women is unknown but could probably protect the infant for at least the neonatal period. As our data show, despite late decrease antibodies levels in breast milk, infants of breastfeeding vaccinated women could be protected for at least six months after vaccination. However, despite observing the passage of antibodies into breast milk, their protective efficacy against COVID-19 in infants and the cut-off for anti-SARS-CoV-2 protection remain unknown. In our cohort, we observed 2 cases of vaccine breakthrough infections in the 6-month follow-up (6%), both in the final study period (between 3 and 6 months). Despite being a small cohort, this represents a higher rate than that observed in previous studies carried out in vaccinated health care professionals^22,23^. Although there are limited data on the incidence of COVID-19 in pregnant women, there are no studies in the literature on SARS-CoV-2 risk of infection in breastfeeding women. Further research is needed to estimate COVID-19 incidence in this group.

We have also observed how, in cases in which a participant became infected by SARS-CoV-2, the levels of IgG-S1 showed a new increase in both serum and breast milk. Not so in the case of IgG-NC which, despite having been positive in serum, did not have its presence in milk confirmed. A possible explanation could be that IgG-NC is a qualitative technique, and the limit of detection could be higher than IG-S1 quantitative antibody determination.

On the basis of the substantial increase in IgG-S1 in serum and milk observed after COVID-19 infection in 2 volunteers, it may be reasonable to consider that a COVID-19 vaccine booster shot could increase serum and milk levels in the same way as a natural infection. AErV observed in our cohort are similar to those demonstrated by the clinical trial conducted for the approval of the vaccine^24^.

The main limitation of this study is its small sample size. In our sample, there was only one infant of exclusive breastfeeding age (<6 months) so it was not possible to analyze whether there is a greater passage of antibodies to milk in this period compared to a longer postpartum period. Moreover, with a larger sample, we could confirm the correlation between antibody levels in serum and breast milk that we found and predict the levels in milk from a serum sample determination.

Furthermore, the use of different techniques for the analysis of the two types of antibodies, IgG-NC and IgG-S1, can lead to a bias in the interpretation of the data.

Most of the studies in pregnant and lactating women to date have been conducted after vaccination with mRNA vaccines. More studies with other vaccines are needed to be able to extrapolate these data to other types of vaccine.

Finally, larger prospective studies are needed to confirm the safety of SARS-CoV-2 vaccination in individuals who are breastfeeding and further assess the association between vaccination and infants’ health and SARS-CoV-2 specific immunity.

## Conclusions

The results of this study show that vaccination with Pfizer-BioNTech is safe during breastfeeding, that it transmits antibodies into breast milk and, in addition, these levels correlate with those observed in serum. IgG serum levels induced by the vaccine decrease over time in a month follow-up, as do the antibodies in breast milk. However, despite late decrease antibody levels in breast milk, infants of breastfeeding vaccinated women could be protected for at least six months after vaccination.

Vaccination against SARS-CoV-2 has been shown to drastically reduce cases of serious infections, as well as the transmissibility of the virus. In breastfeeding mothers this strategy may protect the babies, both by generating group immunity and thus reducing the risk of contracting the infection in their social environment, as well as by the passage of antibodies through breast milk.

## Data Availability

All data produced in the present work are contained in the manuscript

## ACKNOWLEDGMENTS

No compensation was received by any author for their role in this study, and none have conflicts of interest to declare. Only Diaz-Brito and Esteve-Palau had full access to all the data in the study and take responsibility for the integrity of the data and the accuracy of the data analysis.

## AUTHOR CONTRIBUTIONS

Concept and design: Esteve-Palau, Diaz-Brito.

Recruitment of volunteers and laboratory samples collection: Alvarez, Garcia-Terol, Esteve-Palau, García, Moreno and Medina.

Laboratory samples analysis: Gonzalez-Cuevas, Guerrero.

Acquisition, analysis, or interpretation of data: All authors.

Drafting of the manuscript: Esteve-Palau, Casadevall, Diaz-Brito.

Critical revision of the manuscript for important intellectual content: All authors.

Statistical analysis: Esteve-Palau, Casadevall.

Supervision: Diaz-Brito.

